# Antibody response to first and second dose of BNT162b2 in a cohort of characterized healthcare workers

**DOI:** 10.1101/2021.03.24.21254240

**Authors:** Andrea Padoan, Luigi Dall’Olmo, Foscarina della Rocca, Francesco Barbaro, Chiara Cosma, Daniela Basso, Annamaria Cattelan, Vito Cianci, Mario Plebani

**Author notes:** **Corresponding author:** Prof. Mario Plebani, MD, Department of Medicine-DIMED, University of Padova and Department of Laboratory Medicine, University-Hospital of Padova (Italy), Via Giustiniani 2, 35128, Padova, Italy.

## Abstract

**Background:** Vaccine-induced population immunity is a key global strategy to control coronavirus disease 2019 (COVID-19). The rapid implementation and availability of several COVID-19 vaccines is now a global health-care priority but more information about humoral responses to single- and double-dose vaccine is needed

**Methods:** 163 health care workers (HCW) of the Padua University Hospitals, who underwent a complete vaccination campaign with BNT162b2 vaccine were asked to collect serum samples at 12 (t12) and 28 (t28) days after the first inoculum to allow the measurement of SARS-CoV-2 Antibodies (Ab) using chemiluminescent assays against the spike (S) protein and the Receptor Binding Domain (RBD) of the virus, respectively.

**Results:** Significant differences were found at t12 for infection-naïve and subjects with previous-natural infection who present higher values of specific antibodies, while no significant differences have been found between t12 and t28. No statistically significant difference was found between male and female, while lower Ab levels have been observed in subjects older than 60 years at t12 but not at t28.

**Conclusions:** Our study confirms observed differences in vaccine responses between infection-naïve and subjects with previous natural infection at t12 but not for a longer time. The influence of sex and age deserves further studies, even if the relationship with age seems particularly significant.

## Introduction

Coronavirus disease 2019 (COVID-19) is a major public health issue. To contrast the spread of SARS-CoV-2, the rapid immune-induced vaccination has been suggested as a key global strategy and, currently, a total of 82 vaccines are in clinical development, and 182 are in pre-clinical development (https://www.who.int/publications/m/item/draft-landscape-of-covid-19-candidate-vaccines). Several of the vaccines currently developed are based on a double-dose, the prime and boost approach. This strategies allowed to obtain a high immunity, which was demonstrated, for example, to be effective in preventing 95% of Covid-19 cases for BNT162b2 (Pfizer/BioNTech, Comirnaty) [1]. Similarly, strong evidence has been reported that a second dose enhances the antibody response in other vaccines [2,3].

In most European countries, the vaccination campaigns started between the 26 and 31 December 2020 after the first lots of Comirnaty vaccine were delivered [4]. The vaccine generates an immune response against the S1 spike protein, the titers of which Ab correlate with functional viral neutralization [1,5]. Up to now, only few studies have provided pieces of evidence about the immunological status of infection-naïve vaccinated subjects and previous natural infection. Interestingly, very elevated Ab titers were described by two recent studies from Manisty et al. and Prendecki et al, the latter further showing an inverse correlation with age of Ab levels. However, to better clarify immunological response to the two vaccinal inoculums, several time points should be inspected since it has been shown that at least 12-14 days are necessary to mount a valuable antibody response [6]. Furthermore, subjects should be well characterized from their previous infection status. This latter point is particularly important for evaluate the necessity of implementing the boosting dose in previous natural-infection subjects, when Ab levels were already elevated.

Since several immunoassay methods for measuring SARS-CoV-2 Ab were designed to detect N- or generic S-protein antigens, this caused a potential issue on Ab measurement, and methods should be validated for this purpose, or neutralization activity assay should be chosen as alternative. However, this latter technique needs handle live virus in BSL-3 laboratories and for this reason, a surrogate method to evaluate their levels in patients has been strongly advocated [7].

In this study, a well-defined cohort of healthcare workers (HCW) was tested for serum SARS-CoV-2 Ab after 12 and 28 days after the first inoculum of BNT162b2 with two different chemiluminescent immunoassays. The cohort includes a series subjects who presented previous natural infection during the first or second wave of pandemic. The aims of this study are to investigate the Ab levels in previous natural infection and infection-naïve subjects, and to assess further correlations with age and gender.

## Material and Methods

This study included a series of 163 HCW of the Padua University Hospitals, who underwent a complete vaccination campaign (prime dose followed by a boost dose after 21 days) between December 26^th^ 2020 and March 10^th^ 2021. A total of 123 individuals included in this study were previously enrolled in a follow-up study, carried out between April 8 and May 29, 2020, for determining SARS-CoV-2 serological levels as described elsewhere [8], while the other 38 included subjects were post-graduate medical trainees. All subjects underwent periodical nasopharyngeal swab testing (every 2 or 3 weeks) from March 2020 to March 2021. All HCW were asked to collect two serum SARS-CoV-2 S-RBD samples to determining Ab after 12 and 28 days after the first inoculum of BNT162b2; all subjects underwent a second vaccine administration after 21 days from the first dose. For the 38 residents, a pre-vaccination sample was collected from 24 to 0 hours before vaccination. A previous COVID-19 natural infection has been assessed by direct interview and was considered affirmative with at least one positive nasopharyngeal swab test. Serum S-RBD antibodies against the RBD of the Spike (S) protein of the virus was measured by two already validated chemiluminescent immunoassay (CLIA) that determines: a) anti-SARS-CoV-2 S-RBD IgG Ab (Snibe Diagnostics, New Industries Biomedical Engineering Co., Ltd [Snibe], Shenzhen, China), reagent lot 270200211, b) Elecsys Anti-SARS-CoV-2 S (Roche Diagnostics GmbH, Mannheim, Germany), reagent lot 51639401. The anti-SARS-CoV-2 S-RBD IgG Ab assay presents a very good linearity, with a clinical sensitivity and specificity of 96.9% and 91.8%, respectively [9]. The Elecsys Anti-SARS-CoV-2 S, determining total Ab levels, has been extensively evaluated by the manufacturer, showing 99.98 % (95 % CI, 99.91–100 %) specificity on 5,991 samples and 98.8 % (95 % CI, 98.1–99.3 %) sensitivity on 1,423 samples obtained 14 days or later after SARS-CoV-2 PCR-confirmation [10]. Analyses were performed on MAGLUMI™ 2000 Plus (Snibe Diagnostics) and Roche Cobas C8000, and results expressed in kAU/L for both methods. The GraphPad Prism version 9.1 for Windows (GraphPad Software, LLC) and Stata v16.1 (Statacorp, Lakeway Drive, TX, USA) were used for assessing univariate and multivariate differences across the studied groups. The study was performed in accordance with the Declaration of Helsinki and all participants gave informed consent (Institutional Review Board of the University of Padua protocol nr 7862).

## Results

After personal interview, a total of 13 subjects were confirmed to have experienced COVID-19 disease during the first or the second wave of pandemic. Among the 163 individuals, 49 (30.1%) were males and 114 (69.9%) females; age was not different in males and females (Kruskall-Wallis χ^2^=3.783, p = 0.052) and in average was 42.4 years (yrs), standard deviation 11.7 yrs. Considering the group of post-graduate medical trainees with pre-vaccination samples (baseline), 36/38 (94.8%) were infection-naïve. All Ab levels of these 36 individuals were negative (below 1 kAU/L) by Elecsys Anti-SARS-CoV-2 S, while 2/38 (5.2%) were positive for anti-SARS-CoV-2 S-RBD IgG Ab. Notably, one of these subjects is affected by autoimmune disease. On the other hand, among the two subjects with previous-natural SARS-CoV-2 infection, one had negative anti S-RBD levels at baseline for both methods and presented asymptomatic disease. Figure 1, panel A, reports the dot plots of anti S-RBD levels in studied groups. The Snibe anti-SARS-CoV-2 S-RBD IgG median levels for Infection-naïve subjects at 12 days (t 12) were 6.62 kAU/L, 25^th^ and 75^th^ quartiles (IQR) 3.04 and 14.8 kAU/L, respectively, range from 0.56 to 3780 kAU/L, while median levels at 28 days (t28) were 382.0 kAU/L, IQR 141.3 – 727.9 kAU/L and range from 10.7 to 3789 kAU/L. On the same groups, anti S-RBD median levels for Roche at t12 were 2.54 kAU/L, IQR 0.52 – 8.73 kAU/L, range 0.4 to 6316 kAU/L, while median levels at t28 were 1204.0 kAU/L, IQR 510.4 – 2756 kAU/L, range 19.35 kAU/L to 12136 kAU/L. For previous-natural infection disease, Snibe anti-SARS-CoV-2 S-RBD IgG Ab median levels at t12 were 746 kAU/L, IQR 414.7 – 1504.1 kAU/L, range 19.6 to 1504.1 kAU/L, while median levels at t28 were 1713 kAU/L, IQR 870.5 – 2314.5 kAU/L, range 412.5 kAU/L to 8509.3 kAU/L. On the same group, Elecsys Anti-SARS-CoV-2 S at t12 and t28 were all above 12500 kAU/L, being quantifiable only in 3 individuals. Figure 1 shows the Snibe anti-SARS-CoV-2 S-RBD comparative levels at baseline, t12 and t28 for infection-naïve (panel B) and previous-natural infection diseased (panel C). Comparing infection-naïve and previous-natural COVID-19 individuals Snibe anti-SARS-CoV-2 S-RBD values, significant differences were found at t12 for infection-naïve and subjects with previous-natural infection (Kruskall-Wallis test with Bonferroni’s adjusted p-value < 0.001). Furthermore, no significant differences were observable between t12 and t28 of infection-naïve and previous-natural COVID-19 individuals Ab values, respectively (Figure 1, panel D). Further analyses were also performed. Considering anti S-RBD levels and age a significant inverse correlation was found at t12 both for Snibe and Roche (Figure 1, panels E-H). No significant anti S-RBD levels differences were found between males and females in all the studied conditions.

**Figure:**
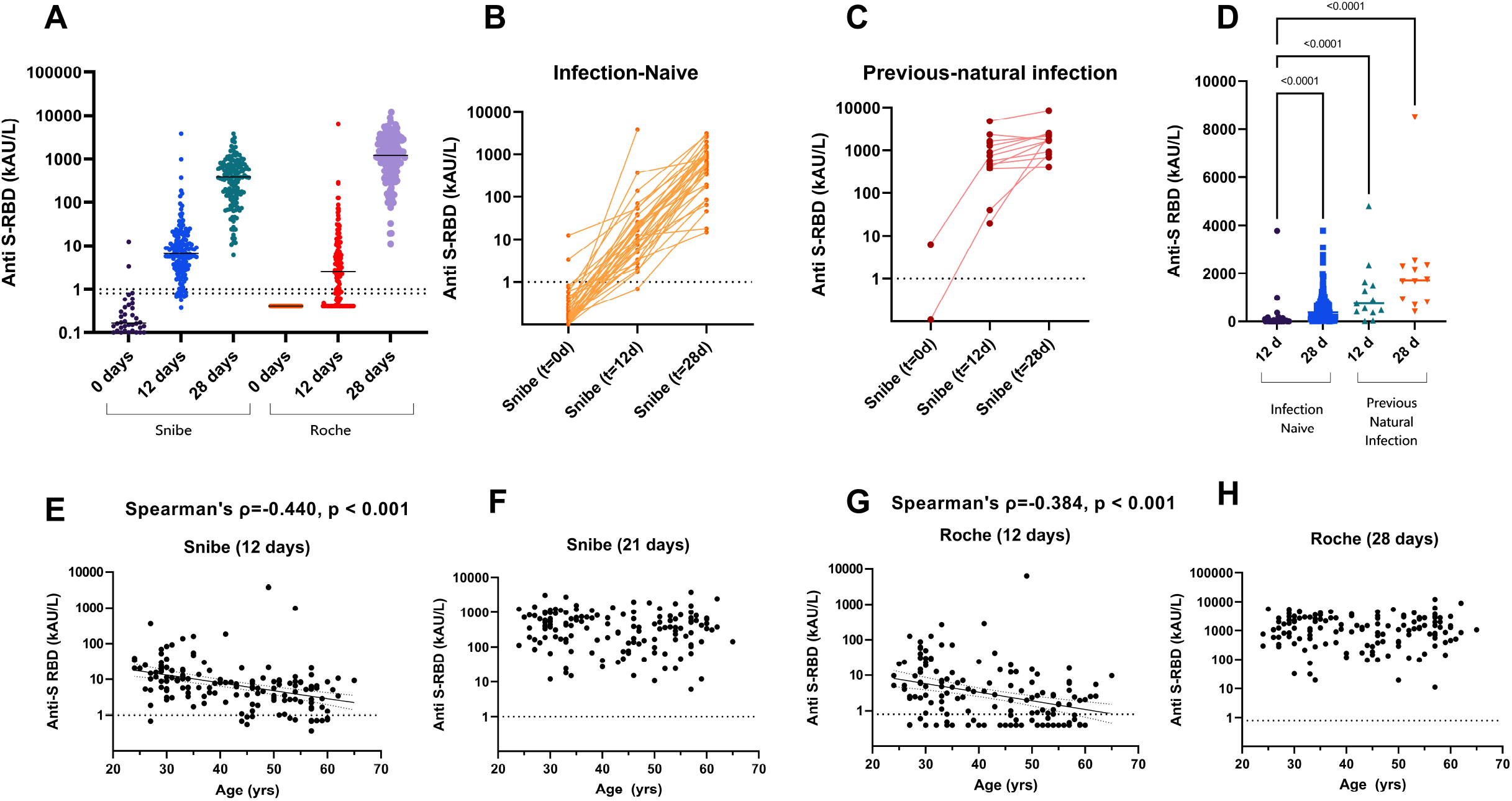
Immunological responses to the two doses of BNT162b2 mRNA vaccine. A) dot-plots of anti S-RBD antibodies measured at baseline (before) and after 12 days or 28 days from the first dose (the dotted lines represent the two cut-offs, 1 kAU/L for Snibe and 0.8 kAU/L for Roche); B-C) comparative anti S-RBD levels at baseline, t12 and t28 (after 12 and 28 days from the first dose) in COVID-19 infection-naïve and previous-natural infection individuals; D) dot-plots of anti S-RBD antibodies measured at t12 and t28 in COVID-19 infection-naïve and previous-natural infection individuals; E - H) correlation of post-vaccination anti-S levels and age with superimposed the linear regression lines (with 95%CI).

## Discussion

Serological studies are a useful tool to estimate the proportion of the population previously infected, to allow the diagnosis of COVID-19 in patients who present late with a low viral load and to assess the efficacy of vaccines in clinical trial [11,12]. Limited data are available on immunoresponses to single- and/or double-dose vaccination, and vaccine responses following previous natural infection have not been assessed in clinical trials [1,5,13]. Recently published papers, however, highlight the value of serological tests before vaccination with SARS-CoV-2 spike messenger RNA vaccines as recipients with preexisting immunity developed systemic side effects at higher frequency than those without preexisting immunity [14]. It should be important, in fact, to avoid reactogenicity after unnecessary boost risks that represent an avoidable and unwelcome in vaccine hesitancy. Significant different antibody responses in infection-naïve individuals and previously infected groups have been reported [15] and further data seem to suggest serological testing as a potential approach to be included at or before the time of first vaccination to prioritize use of boosters doses for individuals with no previous infections [16].

A major issue in evaluating the immune response after vaccination is to use valuable immunoassays which may provide results well correlated with neutralizing antibody titers because they measure both the ability of antibodies to block virus infection and vaccine efficacy [17]. Evidence has been collected to suggest immunoassays using as a target the S1 subunit of the spike protein, the trimeric spike protein or the receptor binding domain of the same spike protein [18]. The results of this study confirm observed differences in vaccine responses between infection-naïve and subjects with previous natural infection at t12 but not for a longer time. The influence of sex and age deserves further studies, even if the relationship with age seems particularly significant.

The paper presents several limitations. First, a more representative number of subjects followed for a longer time is needed. Second, even if anti-S and anti-RBD values have been reported to correlate with in-vitro virus neutralization, the kinetics of neutralizing antibodies after vaccination should be better evaluated. Third, T-cell responses should be evaluated to achieve a better understanding of the immunoresponse after vaccination in both previously infected and infection-naïve subjects. Currently, this is more important as virus variants should affect the efficacy of currently available vaccines and immune-responses mediated by both humoral and T-cells.

## Data Availability

Data will be made available upon request

## Acknowledgments

We thank Giulia Vanuzzo for their valuable technical support. We acknowledge Snibe and Roche diagnostics for kindly supplying reagents without any influence in study design and data analysis.

## References

[1] F.P. Polack, S.J. Thomas, N. Kitchin, J. Absalon, A. Gurtman, S. Lockhart, J.L. Perez, G. Pérez Marc, E.D. Moreira, C. Zerbini, R. Bailey, K.A. Swanson, S. Roychoudhury, K. Koury, P. Li, W. V. Kalina, D. Cooper, R.W. Frenck, L.L. Hammitt, Ö. Türeci, H. Nell, A. Schaefer, S. Ünal, D.B. Tresnan, S. Mather, P.R. Dormitzer, U. Şahin, K.U. Jansen, W.C. Gruber, Safety and Efficacy of the BNT162b2 mRNA Covid-19 Vaccine, N. Engl. J. Med. 383 (2020) 2603–2615.

[2] J.R. Barrett, S. Belij-Rammerstorfer, C. Dold, K.J. Ewer, P.M. Folegatti, C. Gilbride, R. Halkerston, J. Hill, D. Jenkin, L. Stockdale, M.K. Verheul, P.K. Aley, B. Angus, D. Bellamy, E. Berrie, S. Bibi, M. Bittaye, M.W. Carroll, B. Cavell, E.A. Clutterbuck, N. Edwards, A. Flaxman, M. Fuskova, A. Gorringe, B. Hallis, S. Kerridge, A.M. Lawrie, A. Linder, X. Liu, M. Madhavan, R. Makinson, J. Mellors, A. Minassian, M. Moore, Y. Mujadidi, E. Plested, I. Poulton, M.N. Ramasamy, H. Robinson, C.S. Rollier, R. Song, M.D. Snape, R. Tarrant, S. Taylor, K.M. Thomas, M. Voysey, M.E.E. Watson, D. Wright, A.D. Douglas, C.M. Green, A.V.S. Hill, T. Lambe, S. Gilbert, A.J. Pollard, Phase 1/2 trial of SARS-CoV-2 vaccine ChAdOx1 nCoV-19 with a booster dose induces multifunctional antibody responses, Nat. Med. 27 (2021) 279–288.

[3] E.J. Anderson, N.G. Rouphael, A.T. Widge, L.A. Jackson, P.C. Roberts, M. Makhene, J.D. Chappell, M.R. Denison, L.J. Stevens, A.J. Pruijssers, A.B. McDermott, B. Flach, B.C. Lin, N.A. Doria-Rose, S. O’Dell, S.D. Schmidt, K.S. Corbett, P.A. Swanson, M. Padilla, K.M. Neuzil, H. Bennett, B. Leav, M. Makowski, J. Albert, K. Cross, V.V. Edara, K. Floyd, M.S. Suthar, D.R. Martinez, R. Baric, W. Buchanan, C.J. Luke, V.K. Phadke, C.A. Rostad, J.E. Ledgerwood, B.S. Graham, J.H. Beigel, Safety and Immunogenicity of SARS-CoV-2 mRNA-1273 Vaccine in Older Adults, N. Engl. J. Med. 383 (2020)

[4] European center for disease prevention and control (ECDC), Overview of COVID-19 vaccination strategies and vaccine deployment plans in the EU / EEA and the UK Key findings, (2020) 1–22.

[5] E.E. Walsh, R.W. Frenck, A.R. Falsey, N. Kitchin, J. Absalon, A. Gurtman, S. Lockhart, K. Neuzil, M.J. Mulligan, R. Bailey, K.A. Swanson, P. Li, K. Koury, W. Kalina, D. Cooper, C. Fontes-Garfias, P.-Y. Shi, Ö. Türeci, K.R. Tompkins, K.E. Lyke, V. Raabe, P.R. Dormitzer, K.U. Jansen, U. Şahin, W.C. Gruber, Safety and Immunogenicity of Two RNA-Based Covid-19 Vaccine Candidates, N. Engl. J. Med. 383 (2020) 2439–2450.

[6] M. Plebani, A. Padoan, D. Negrini, B. Carpinteri, L. Sciacovelli, Diagnostic performances and thresholds: the key to harmonization in serological SARS-CoV-2 assays?, Clin. Chim. Acta. 509 (2020) 1–7.

[7] F. Amanat, D. Stadlbauer, S. Strohmeier, T.H.O. Nguyen, V. Chromikova, M. McMahon, K. Jiang, G.A. Arunkumar, D. Jurczyszak, J. Polanco, M. Bermudez-Gonzalez, G. Kleiner, T. Aydillo, L. Miorin, D.S. Fierer, L.A. Lugo, E.M. Kojic, J. Stoever, S.T.H. Liu, C. Cunningham-Rundles, P.L. Felgner, T. Moran, A. García-Sastre, D. Caplivski, A.C. Cheng, K. Kedzierska, O. Vapalahti, J.M. Hepojoki, V. Simon, F. Krammer, A serological assay to detect SARS-CoV-2 seroconversion in humans, Nat. Med. 26 (2020) 1033–1036.

[8] F. Barbaro, F. Della Rocca, A. Padoan, A. Aita, V. Cianci, D. Basso, A. Cattelan, D. Donato, M. Plebani, L. Dall’Olmo, A longitudinal study of healthcare workers’ surveillance during the ongoing COVID-19 Epidemics in Italy: is SARS-CoV-2 still a threat for the Health-care System?, MedRxiv. (2021) 2021.02.23.21249481. https://doi.org/10.1101/2021.02.23.21249481.

[9] Padoan, F. Bonfante, C. Cosma, C. Di Chiara, L. Sciacovelli, M. Pagliari, A. Bortolami, P. Costenaro, G. Musso, D. Basso, C. Giaquinto, M. Plebani, Analytical and clinical performances of a SARS-CoV-2 S-RBD IgG assay: comparison with neutralization titers, MedRxiv. (2021) 2021.03.10.21253260. https://doi.org/10.1101/2021.03.10.21253260.

[10] M. Poljak, A. Oštrbenk Valenčak, T. Štamol, K. Seme, Head-to-head comparison of two rapid high-throughput automated electrochemiluminescence immunoassays targeting total antibodies to the SARS-CoV-2 nucleoprotein and spike protein receptor binding domain, J. Clin. Virol. 137 (2021) 104784.

[11] M. Plebani, SARS-CoV-2 antibody-based SURVEILLANCE: New light in the SHADOW, EBioMedicine. 61 (2020) 103087.

[12] R. Li, S. Pei, B. Chen, Y. Song, T. Zhang, W. Yang, J. Shaman, Substantial undocumented infection facilitates the rapid dissemination of novel coronavirus (SARS-CoV-2), Science (80). 368 (2020)

[13] L.R. Baden, H.M. El Sahly, B. Essink, K. Kotloff, S. Frey, R. Novak, D. Diemert, S.A. Spector, N. Rouphael, C.B. Creech, J. McGettigan, S. Khetan, N. Segall, J. Solis, A. Brosz, C. Fierro, H. Schwartz, K. Neuzil, L. Corey, P. Gilbert, H. Janes, D. Follmann, M. Marovich J. Mascola, L. Polakowski, J. Ledgerwood, B.S. Graham, H. Bennett, R. Pajon, C. Knightly, B. Leav, W. Deng, H. Zhou, S. Han, M. Ivarsson, J. Miller, T. Zaks, Efficacy and Safety of the mRNA-1273 SARS-CoV-2 Vaccine, N. Engl. J. Med. 384 (2021) 403–416.

[14] F. Krammer, K. Srivastava, H. Alshammary, A.A. Amoako, M.H. Awawda, K.F. Beach, M.C. Bermúdez-González, D.A. Bielak, J.M. Carreño, R.L. Chernet, L.Q. Eaker, E.D. Ferreri, D.L. Floda, C.R. Gleason, J.Z. Hamburger, K. Jiang, G. Kleiner, D. Jurczyszak, J.C. Matthews, W.A. Mendez, I. Nabeel, L.C.F. Mulder, A.J. Raskin, K.T. Russo, A.-B.T. Salimbangon, M. Saksena, A.S. Shin, G. Singh, L.A. Sominsky, D. Stadlbauer, A. Wajnberg V. Simon, Antibody Responses in Seropositive Persons after a Single Dose of SARS-CoV-2 mRNA Vaccine., N. Engl. J. Med. (2021) NEJMc2101667.

[15] M. Prendecki, C. Clarke, J. Brown, A. Cox, S. Gleeson, M. Guckian, P. Randell, A.D. Pria, L. Lightstone, X.-N. Xu, W. Barclay, S.P. McAdoo, P. Kelleher, M. Willicombe, Effect of previous SARS-CoV-2 infection on humoral and T-cell responses to single-dose BNT162b2 vaccine., Lancet. 6736 (2021) 10–12.

[16] C. Manisty, A.D. Otter, T.A. Treibel, ine McKnight, D.M. Altmann, T. Brooks, M. Noursadeghi, R.J. Boyton, A. Semper, J.C. Moon, Correspondence Antibody response to first BNT162b2 dose in previously SARS-CoV-2-infected individuals, Lancet. 6736 (2021) 2–3.

[17] V.V. Edara, W.H. Hudson, X. Xie, R. Ahmed, M.S. Suthar, Neutralizing Antibodies Against SARS-CoV-2 Variants After Infection and Vaccination., JAMA. (2021). doi:10.1001/jama.2021.4388.

[18] F. Bonelli, F.A. Blocki, T. Bunnell, E. Chu, O.A. De La, D.G. Grenache, G. Marzucchi, E. Montomoli, L. Okoye, L. Pallavicini, V.A. Streva, A. Torelli, A. Wagner, D. Zanin, C. Zierold, J.J. Wassenberg. Evaluation of the automated LIAISON® SARS-CoV-2 TrimericS IgG assay for the detection of circulating antibodies. Clin Chem Lab Med. 2021 [ahead of print]. doi: 10.1515/cclm-2021-0023.

